# Folate Overload and the Placental Hormone Axis: A Hidden Risk for Gestational Diabetes Mellitus

**DOI:** 10.1101/2025.06.05.25329035

**Authors:** Tanja Jankovic-Karasoulos, Melanie D Smith, Shalem Leemaqz, Murthy Mittinty, Jessica Williamson, Dylan McCullough, Anya L Arthurs, Gustaaf A Dekker, Claire T Roberts

## Abstract

**Background:** The prevalence of gestational diabetes mellitus (GDM) in Australia has increased more than threefold, from 5.6% in 2010 to 19.3% in 2022. The onset of the sharp upward trend coincides with the introduction of mandatory folic acid (FA) food fortification and increased FA supplementation during pregnancy. Animal studies provide evidence that high FA intake in pregnancy can induce insulin resistance and hyperglycaemia in the mother and/or the offspring, but the underlying mechanisms are unknown.

**Objective:** To determine whether high FA intake has altered maternal folate status in a manner that increases GDM risk, and whether key hormones that regulate maternal glucose homeostasis are affected following fortification.

**Methods:** We measured serum folate, red cell folate (RCF), prolactin (PRL), human placental lactogen (hPL) and placental growth hormone (GH2) in early pregnancy blood samples collected from women enrolled in two prospective cohorts: SCOPE (N=1164) recruited before the FA fortification mandate and STOP (N=1300) recruited after its implementation.

**Results:** Compared to women pre-fortification, women post fortification had a higher incidence of GDM (5.0% vs 15.2%), and elevated levels of serum folate (↑18%), RCF (↑259%), hPL (↑29%), and GH2 (↑13%). RCF concentrations above the clinical reference range were found in 57.6% of women post fortification. Causal mediation analysis suggests that higher RCF contributed to increased GDM risk in this group. Compared to women with RCF concentrations within the reference range, those with RCF excess (defined as concentrations well above this range) had 48% more GDM cases, and significantly higher PRL (↑24.2%) and hPL (↑12.7%) concentrations.

**Conclusions:** Maternal folate excess is likely contributing to the rising prevalence of GDM in Australia. These findings highlight the need to evaluate excess FA/folate safety in pregnancy, particularly in countries with mandatory fortification. Placental hormones may represent a mechanistic link between excess folate and GDM, warranting further investigation.

## Introduction

According to national surveillance data, gestational diabetes mellitus (GDM) incidence has more than tripled in Australia in just over a decade (Figure 1) (1). The most recent change in GDM diagnostic criteria by the World Health Organization (WHO) (2) was implemented in Australia in 2015. While the diagnostic change has often been cited to account for the rising GDM rates, the steep increase in incidence both predates its introduction and continues thereafter (Figure 1).

**Figure 1.**
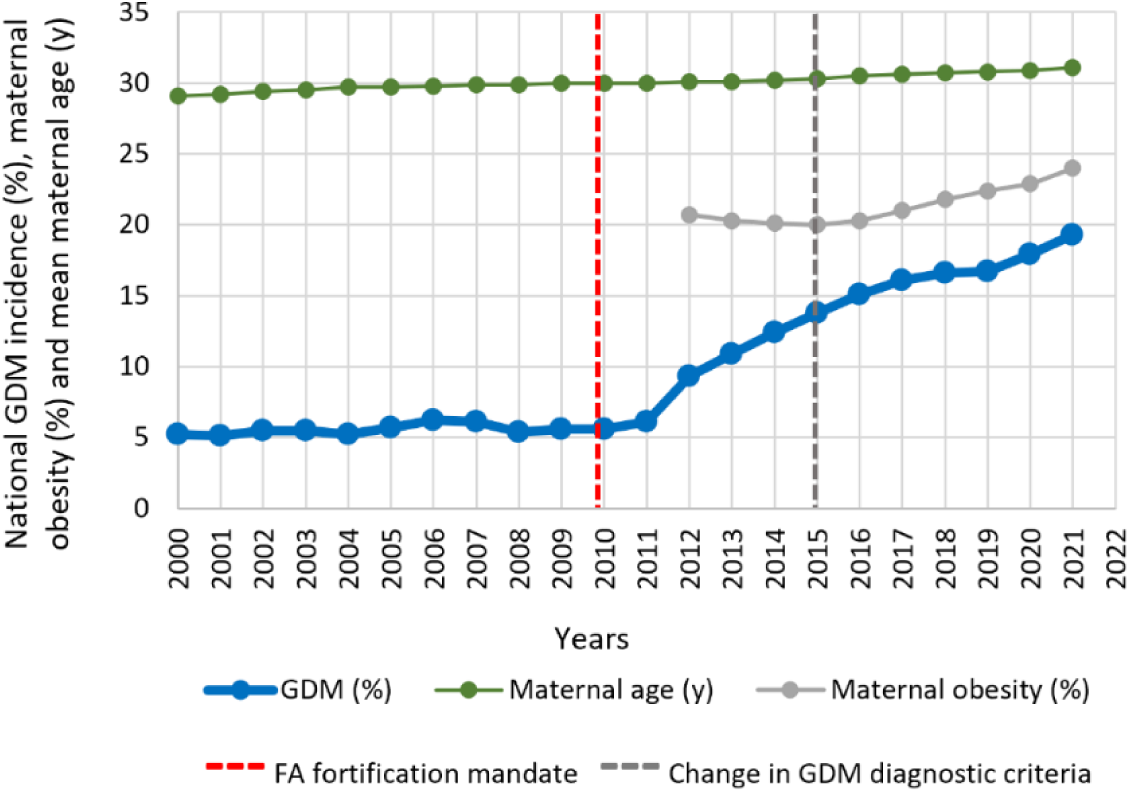
Incidence of GDM, maternal age and obesity in Australia. (AIHW trends over time data) (1). National GDM incidence is indicated by the solid blue line; Mean national maternal age is shown in green; National maternal obesity data (grey) is incomplete prior to 2012; Dashed vertical lines indicate the implementation of the FA food fortification mandate (red) and the new WHO GDM diagnostic criteria (dark grey).

Well established GDM risk factors such as maternal obesity and age (Figure 1) as well as ethnicity, have marginally changed during the same period. Although these factors may have contributed toward the GDM rise, even combined they cannot explain the observed trajectory. Moreover, the same risk factors also increase the risk for hypertensive disorders of pregnancy which are not on the same trajectory as GDM. This suggests that additional factors are involved.

Observational studies link high folic acid (FA; synthetic folate) intake with increased GDM risk (3–6), while animal studies provide compelling evidence that high FA intake during pregnancy plays a role in insulin resistance and impaired glucose handling (7–10), but the mechanisms remain unknown. The WHO recommends women supplement with 400 µg FA daily when trying to conceive until 12 weeks’ gestation, to reduce the risk of foetal neural tube defects (11). Accordingly, FA supplementation with a daily dose of 400µg has been part of the Australian clinical pregnancy guidelines for more than 20 years. However, given the neural tube closes ∼ 4 weeks post-conception (12), before most women know they are pregnant, and that many pregnancies are unplanned, in 2009 the government mandated FA food fortification. Concurrent with this has been an increase in the use of FA containing prenatal multivitamins, with some leading brands containing double the recommended FA dose (800 μg). Our recent study shows that the period spanning FA food fortification mandate was accompanied by an increase in the use of supplements that contain 800 µg or more FA, from 24% pre fortification to over 60% post fortification (13). A recent systematic review of women taking FA supplements in countries with mandatory food fortification programs, including Australia, found that almost all women exceed 1000 μg daily limit (14). Whilst this limit is set due to potential masking of B12 deficiency, to date there has been no established upper limit for adverse pregnancy outcomes. Importantly, recent studies report an increase in women who continue to take FA containing multivitamins throughout pregnancy (15), despite no known benefit or established harm from continuing to supplement beyond 12 weeks, long after the neural tube has closed. Increased dose and duration of FA supplementation, and high levels of unmetabolized FA and folate in maternal blood during pregnancy, have raised concerns about potential harms (16) (6).

To determine whether increased FA intake over the past decade has placed Australian women at increased risk of GDM, we leveraged two large prospective pregnancy cohorts that were recruited at the same hospital prior to (SCOPE), and post (STOP), the 2009 FA fortification mandate. We set out to determine the incidence of GDM in these cohorts to ensure that our local data reflects the broader population trend, and to measure maternal folate status to establish risk. In search of a mechanism by which FA may influence GDM risk, we turned to the placenta, an organ unique to pregnancy that plays a central role in regulating maternal glucose homeostasis. The placenta secretes hormones into the maternal circulation such as placental growth hormone variant (GH2) and human placental lactogen (hPL) which, along with prolactin (PRL) released from the maternal pituitary and uterine decidua, modulate maternal insulin sensitivity and glucose homeostasis (17) (18). These hormones contribute to the physiological insulin resistance of pregnancy, a critical adaptation that increases maternal glucose availability for the foetus, while also promoting compensatory insulin secretion to maintain maternal glucose homeostasis and prevent hyperglycaemia and GDM (17).

Our hypothesis that FA may exert its effects via the placenta is further supported by epidemiological data: while GDM incidence has increased markedly, the prevalence of Type 2 diabetes - despite its pathophysiological similarities to GDM - has remained relatively stable in Australia over the same period (19). This divergence suggests that the rising burden of GDM is not simply a reflection of general metabolic trends and reinforces the possibility that FA exerts pregnancy-specific effects on glucose metabolism, potentially mediated through placental endocrine function.

Given that placental function is highly reliant on maternal nutrient supply (20), and that FA/folates play a key role in epigenetic regulation of gene expression and protein synthesis (21), it is plausible that increased FA/folate supply from the mother to the placenta can alter placental function (hormone secretion) placing women at increased risk of GDM. We aimed to determine whether high maternal folate increases the risk for GDM in women post FA fortification, and whether placental hormones that regulate maternal glucose homeostasis are involved.

## Research Design and Methods

### Study cohorts and criteria

We used data from two prospective pregnancy cohorts that recruited women at the Lyell McEwin Hospital, Adelaide, South Australia. The Adelaide branch of the international SCOPE (<u>Sc</u>reening f<u>o</u>r <u>P</u>regnancy <u>E</u>ndpoints) cohort recruited nulliparous women with singleton pregnancies prior to the FA fortification mandate (2005-2008; Total N=1164), while the STOP (<u>S</u>creening <u>T</u>ests to identify poor <u>O</u>utcomes of <u>P</u>regnancy) cohort was recruited 6-9 years post fortification mandate (2015-2018; Total N=1300). Women at high risk of pregnancy complications due to underlying medical history (e.g. type 1 or type 2 diabetes, hypertension, and related disorders), 3 or more previous miscarriages or terminations of pregnancy, or those whose pregnancy was complicated by a known foetal anomaly or if they received interventions that may modify pregnancy outcome (e.g. aspirin) were excluded from both studies. Women were also excluded if they were taking calcium (>1 g/d), eicosapentaenoic acid (≥2.7 g/d), vitamin C (>1,000 mg/d) or vitamin E (>400 IU/d). Only women with a confirmed GDM pregnancy outcome were included in data analyses. Importantly, SCOPE and STOP GDM incidence was calculated using the same WHO diagnostic criteria (2). Non-GDM cases include uncomplicated pregnancies as well as pregnancies affected by pathologies such as Gestational hypertension (GHTN), Preeclampsia (PE), Spontaneous Preterm Birth (sPTB) and Small for Gestational Age (SGA). SCOPE and STOP study ethics approvals were obtained from the Central Northern Adelaide Health Service Human Research Ethics Committee (REC1712/5/2008) and the Women’s and Children’s Health Network Human Research Ethics Committee (HREC/14/WCHN/90), respectively. SCOPE and STOP were registered with the Australian New Zealand Clinical Trial Registry (ACTRN12607000551493 and ACTRN12614000985684, respectively).

### Blood biochemical measurements

Peripheral non-fasting blood samples collected at 14-16 (SCOPE; Mean gestational age 15 weeks’) and 6-16 (STOP; Mean gestational age 12 weeks’) weeks of gestation were placed on ice, processed, and stored at -80°C for future analyses. Serum folate, red cell folate (RCF), PRL, hPL and GH2 were measured as described previously (13). Sample numbers available for each hormone analysis are presented in Figure 2. RCF measurements were available for 1191 women in the post FA fortification STOP cohort but not for SCOPE. However, we previously published RCF data from 410 women at 12 weeks’ gestation who were recruited at the same hospital and at the same time as SCOPE women, and thus prior to FA fortification mandate (13). This enabled us to compare changes in RCF in pregnancies pre and post FA fortification mandate.

**Figure 2.**
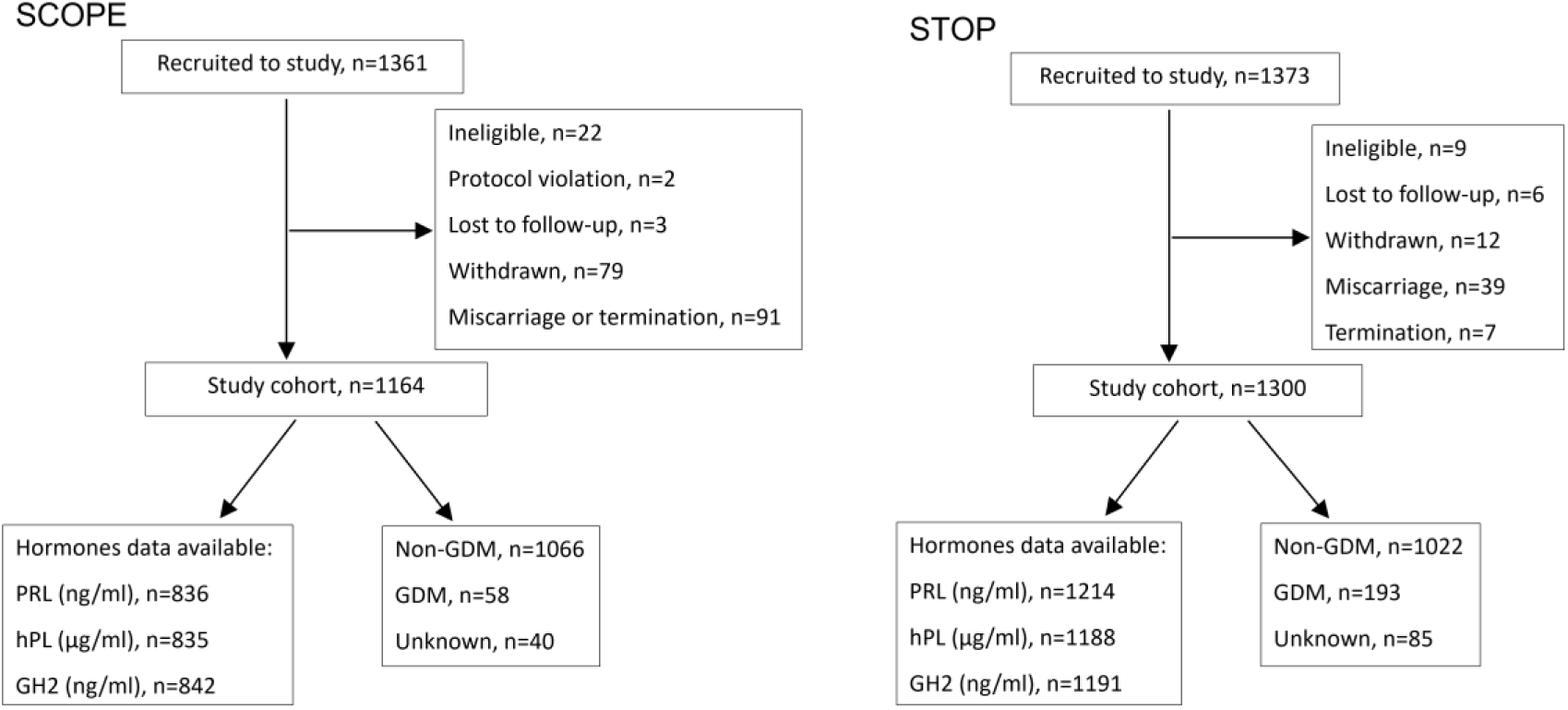
Flow diagram for SCOPE and STOP studies showing final sample numbers available for hormone measurements.

### Statistical methodology

Descriptive statistics for maternal characteristics are reported by cohort. We conducted an individual participant data meta-analysis to allow for the case-mix heterogeneity (arising when the intervention effect is modified by one or more of the confounders used in defining the case-mix) and “beyond case-mix heterogeneity” (due to study differences based on design or methodological aspects). Following the work of Vo *et al*. (2019) (22), we performed a meta-analysis involving two cohorts (SCOPE and STOP) to compare the effect of serum folate concentration as continuous exposure on a dichotomous outcome GDM. As each participant has only one observed outcome, a counterfactual outcome (if the participant was assigned to a different folate exposure) is calculated. To adjust for observed confounding, inverse probability treatment weights (IPTW) were obtained for the continuous exposure (serum and red cell folate concentrations) with maternal BMI, age, ethnicity, SEI, and metabolic syndrome as covariates within each study, where a normal probability density is assumed. Due to other epidemiological factors that may induce differences between the cohorts, the version of folate exposure is likely to be different between cohorts, hence a propensity of study was estimated using Logistic regression with the same covariates. An overall weight was obtained by multiplying the treatment weights with the propensity of study. To estimate relative risk and corresponding 95% confidence intervals we used log-binomial regression to fit the outcome model, with serum folate as the exposure and GDM as the outcome, weighted by the overall weight, we also used study as confounder. The IPTW analysis was conducted under the assumptions of consistency, positivity, and ignorable treatment (Supplemental Material). To analyse the trend of hormone concentration across gestation, linear model with cluster-robust Sandwich variance was used. This allows for prediction of the mean hormone level at 16 weeks’ gestation, adjusted for the gestational age at sampling. Hormone levels were log-transformed to approximate normality, and cohort, gestational age, and their interaction term included as predictors. Causal mediation analysis (which allows inference of causation) was performed to estimate the direct effect of red cell folate on the odds of GDM and the joint indirect effect of red cell folate on GDM through hPL and GH2. The natural direct and indirect effects were obtained using the imputation approach (23) and medflex R package (24). All statistical analyses were performed using R version 4.3.2 (R Foundation for Statistical Computing, Vienna, Austria).

## Results

### Comparative assessment of maternal factors and pregnancy outcomes in SCOPE and STOP

Maternal characteristics and pregnancy outcome data for 1164 SCOPE, and 1300 STOP pregnancies are presented in Table 1. STOP data are compared to SCOPE throughout this section.

**Table 1.** Maternal characteristics assessed in early gestation in the SCOPE (15 weeks’) and STOP (12 weeks’) cohorts with available term pregnancy outcome data.

|  | SCOPE<br>(N=1164) | STOP<br>(N=1300) | P-value |
| --- | --- | --- | --- |
| Maternal age (y): Median (IQ range) | 23.0 (20.0-27.0) | 26.0 (22.0-29.0) | <0.0001 |
| Maternal BMI: Median (IQ range) | 25.6 (22.2-30.8) | 26.3 (22.7-31.8) | 0.006 |
| Maternal BMI (category): N(%) |  |  | 0.02 |
| Missing | 0 (0.0) | 1 (0.1) |  |
| <20 | 122 (10.5) | 95 (7.3) |  |
| ≥20 to <25 | 398 (34.2) | 442 (34.0) |  |
| ≥25 to <30 | 322 (27.7) | 361 (27.8) |  |
| ≥30 to <40 | 267 (22.9) | 315 (24.2) |  |
| ≥40 | 55 (4.7) | 86 (6.6) |  |
| SEI*: Median (IQ range) | 25.0 (20.0-30.0) | 29.0 (22.0-45.0) | <0.0001 |
| Ethnicity: Caucasian N(%) |  |  | 0.0005 |
| Yes | 1067 (91.7) | 1073 (82.5) |  |
| No | 97 (8.3) | 227 (17.5) |  |
| Smoking: N(%) |  |  | <0.0001 |
| Missing | 0 (0.0) | 7 (0.5) |  |
| No | 886 (76.1) | 1077 (82.8) |  |
| Yes | 278 (23.9) | 216 (16.6) |  |
| Folic acid supplementation (µg/day): N(%) |  |  | <0.0001 |
| Missing | 0 (0.0) | 152 (11.7) |  |
| None | 251 (21.6) | 97 (7.5) |  |
| ≤400 | 116 (10.0) | 15 (1.2) |  |
| >400 to <800 | 495 (42.5) | 256 (19.7) |  |
| ≥800 | 302 (25.9) | 780 (60.0) |  |
| Metabolic Syndrome: N(%) |  |  | <0.0001 |
| Missing | 11 (0.9) | 92 (7.1) |  |
| No | 953 (81.9) | 1096 (84.3) |  |
| Yes | 200 (17.2) | 112 (8.6) |  |
| Pregnancy Outcome |  |  |  |
| Gestational diabetes mellitus (GDM)† | 58 (5.0) | 198 (15.2) | <0.0001 |
| Gestational hypertension (GHTN) | 94 (8.1) | 83 (6.4) | 0.14 |
| Preeclampsia (PE) | 117 (10.1) | 120 (9.2) | 0.37 |
| Spontaneous preterm birth (sPTB) | 69 (5.9) | 61 (4.7) | 0.14 |
| Small for gestational age (SGA)‡ | 141 (12.1) | 155 (11.9) | 0.65 |

\*SEI = New Zealand Socio-economic Index, a scale of 10-90 with a lower score indicating greater disadvantage;
†SCOPE and STOP GDM incidence were calculated using the same 2014 diagnostic criteria (old criteria applied at the time of SCOPE (2005-2008) had SCOPE GDM incidence at 4.4%); ‡SGA defined as birthweight below the 10<sup>th</sup> customised centile adjusted for maternal height, weight, parity, ethnicity, gestational age at delivery and infant sex.

STOP women were older, had a higher BMI, SEI (but still disadvantaged), were less likely to be Caucasian and less likely to smoke cigarettes (Table 1).

At least 60% of STOP women reported daily FA supplementation of 800μg or more, compared to 25.9% in SCOPE (Table 1). The magnitude of this difference is likely an underestimation due to 11.7% missing supplementation data in STOP. Folic acid supplementation categories were based on the recommended daily supplementation dose in pregnancy (400μg) and the commonly used higher dose of 800μg found in many prenatal supplements.

Fewer STOP women met the criteria for metabolic syndrome, although some data were missing (7.1%; Table 1). Importantly, even if all missing cases were positive, the incidence would remain lower than in SCOPE. This is important given that metabolic syndrome is an independent risk factor for GDM (25).

All assessed pregnancy outcomes were comparable between cohorts, except for GDM (Table 1), which showed a marked increase in the STOP cohort (Table 1). The incidence of GDM had more than tripled, aligning with the national rise over the same period (Figure 1). Importantly, this increase remained significant after adjusting for established risk factors including maternal age, BMI, ethnicity, SEI and metabolic syndrome.

### Maternal folate status pre and post FA fortification

Serum folate concentrations reflect recent intake and must be interpreted with caution in non-fasting samples. Nevertheless, compared to women pre- FA fortification, women post FA fortification had 18% higher serum folate (Median [IQ range]: 36 [28.2-2.8] vs 40.5 [36.3-45.3] nmol/L, p<0.0001).

Although some health authorities recommend measuring serum folate for diagnosis of folate deficiency, red cell folate (RCF) concentration is a more reliable indicator of long-term folate stores. Importantly, RCF concentration is a surrogate marker of tissue folate levels (26). Compared to women pre- FA fortification, RCF was 259% higher in women post FA fortification (Median [IQ range]: 561 [370-820.5] vs 1490 [1222.5-1787.5] nmol/L, p<0.0001; Figure 3). This difference is likely underestimated as a large proportion of post FA fortification samples reached the upper detection limit of the assay (1790 nmol/L; Figure 3).

**Figure 3:**
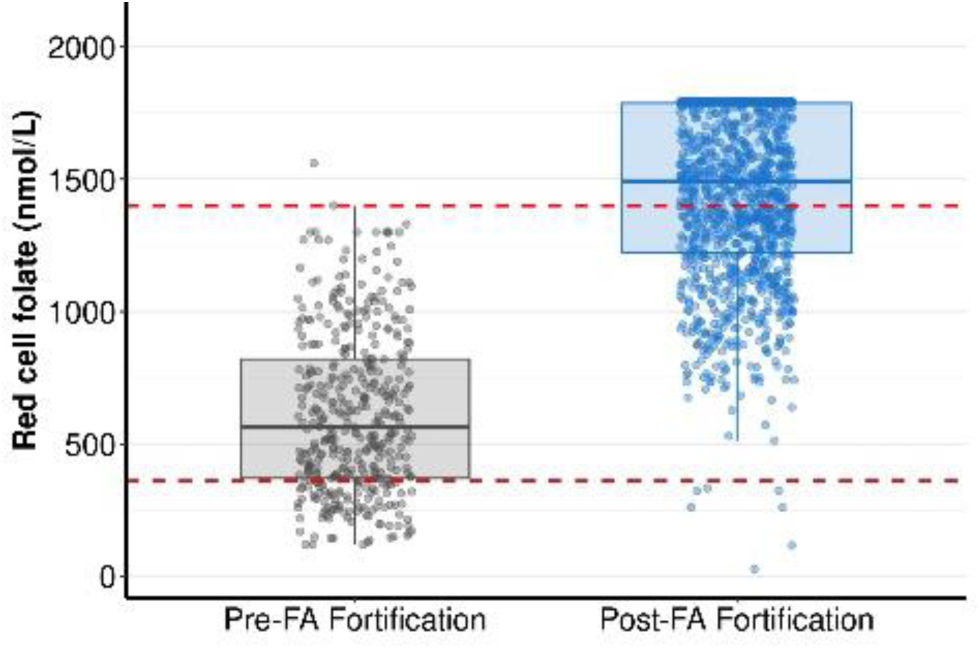
Red cell folate (RCF) concentration in pregnant women prior to and post FA food fortification. Data in the pre- and post- FA fortification categories are presented as median (IQ range). Red dotted lines indicate the normal reference range for RCF (360-1400 nmol/L) set by the Royal College of Pathologists of Australasia (RCPA). Assay upper limit of detection is 1790 nmol/L. RCF measures were compared between STOP (post-FA fortification; using N=1191 samples for which we had available RCF data) and a separate pre-FA fortification cohort (not SCOPE, N=410) using Mann-Whitney U test.

The *normal* reference range for red cell folate is 360 - 1400 nmol/L. In this study, RCF levels below this range are referred to as *deficient*, levels above the reference range but below the assay’s upper detection limit (1790 nmol/L) as *elevated*, and levels at or above the detection limit as *excess*. The proportion of women with RCF deficiency declined from 22.7% (N=93) pre- FA fortification to just 0.6% (N=7) post FA fortification (Figure 3). In contrast, the proportion of women with RCF levels above the normal reference range increased markedly from 0.5% (N=2) pre- FA fortification to 57.6% (N=686) post FA fortification (Figure 3). These findings suggest that, in the post fortification era, elevated and excess maternal folate levels have largely replaced deficiency.

### Red cell folate increases GDM risk in women post-fortification

An inverse probability weighting model was used to assess the overall relative risk of developing GDM in STOP compared to SCOPE. After adjusting for established GDM risk factors (maternal age, BMI, ethnicity, SEI, and metabolic syndrome), serum folate alone did not significantly increase GDM risk (Table 2). However, a combination of serum folate and study (i.e., post-fortification) placed women post FA fortification at greater than three-fold increased risk of developing GDM compared to women pre- FA fortification (Table 2). This risk estimate is consistent with the measured three-fold increased GDM incidence in STOP compared to SCOPE (Table 1).

**Table 2.**
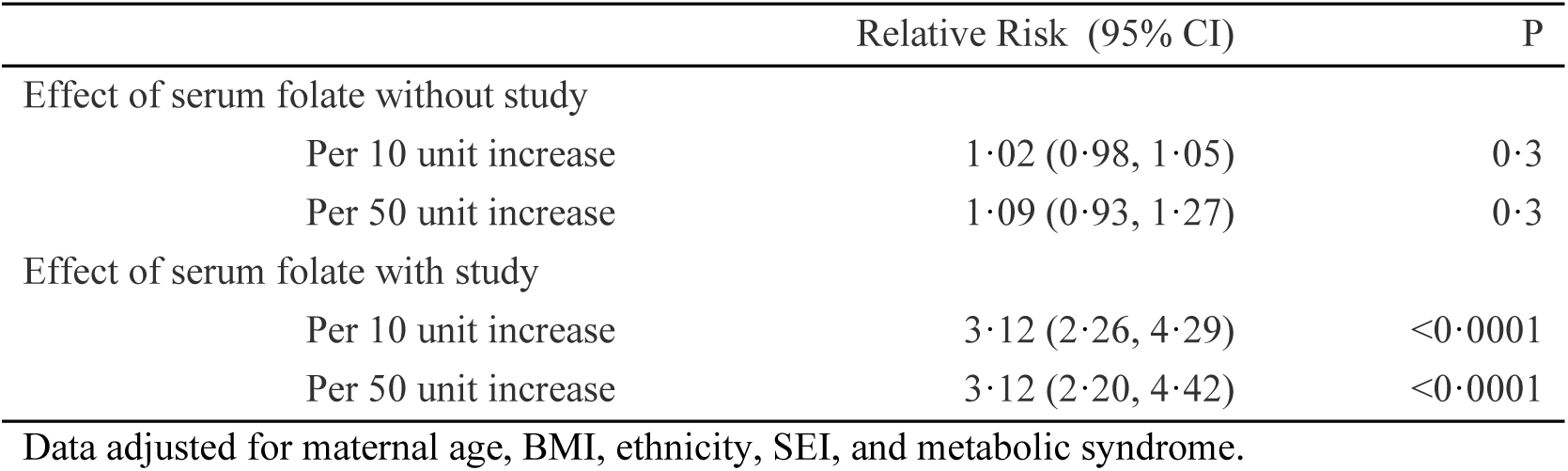
The effect of serum folate and study (STOP vs SCOPE) on GDM risk.

|  | Relative Risk (95% CI) | P |
| --- | --- | --- |
| Effect of serum folate without study |  |  |
| Per 10 unit increase | 1.02 (0.98, 1.05) | 0.3 |
| Per 50 unit increase | 1.09 (0.93, 1.27) | 0.3 |
| Effect of serum folate with study |  |  |
| Per 10 unit increase | 3.12 (2.26, 4.29) | <0.0001 |
| Per 50 unit increase | 3.12 (2.20, 4.42) | <0.0001 |

Data adjusted for maternal age, BMI, ethnicity, SEI, and metabolic syndrome.

As RCF data were not available for SCOPE, we could not directly assess whether the 259% increase in RCF post FA fortification accounted for the cohort effect on GDM risk. However, when we analysed the STOP data alone, to determine what factors increased GDM risk *within* the cohort, we found that both triglycerides and RCF significantly increased the relative risk of GDM in STOP women after adjusting data for known GDM risk factors including maternal age, BMI, ethnicity, SEI, and metabolic syndrome. For every 0.5 unit increase in triglycerides the risk of developing GDM increased by 28% (Table 3). Whilst triglycerides are an independent GDM risk factor in STOP, they are unlikely to explain the increased GDM incidence in STOP compared to SCOPE as a similar relative risk was observed in SCOPE (Relative risk for 0.5 unit increase in triglycerides in SCOPE [95% CI]: 1.34 [1.2-1.5], p<0.0001), and median triglyceride levels were lower in STOP compared to SCOPE (Median [IQR] 1.1 [0.9-1.4] mmol/L vs 1.4 [1.1-1.8] mmol/L, p<0.0001, respectively).

**Table 3.** Factors contributing to GDM risk in women post FA fortification (STOP)

|  | Relative Risk (95% CI) | P |
| --- | --- | --- |
| Effect of triglycerides |  |  |
| Per 0.1 unit increase | 1.05 (1.03, 1.07) | <0.0001 |
| Per 0.5 unit increase | 1.28 (1.14, 1.43) | <0.0001 |
| Effect of red cell folate (RCF) |  |  |
| Per 100 unit increase | 1.06 (1.00, 1.12) | 0.04 |
| Per 500 unit increase | 1.34 (1.01, 1.78) | 0.04 |

Data adjusted for maternal age, BMI, ethnicity, SEI, and metabolic syndrome.

In contrast, changes in RCF could explain the increase in GDM incidence in STOP. For every 500 nmol/L increase in RCF the risk of developing GDM increased by 34% (Table 3). Given the rise in median RCF from 561 [370-820.5] nmol/L pre- FA fortification to 1490 [1223-1788] nmol/L post FA fortification (Median [IQ range]; p<0.0001; Figure 3), this substantial increase in folate status may plausibly contribute to increased GDM risk and higher GDM incidence in STOP compared to SCOPE.

### Red cell folate strata reflect a stepwise rise in GDM incidence in women post fortification

To support the risk prediction findings that higher RCF increases GDM risk in women post FA fortification, we stratified the measured GDM incidence in STOP by red cell folate status (using RCF data presented in Figure 3). Findings are presented in Table 4.

**Table 4.** GDM incidence stratified by RCF category among 1191 STOP women with available RCF data.

| RCF Reference Range | Total Number of Women (N) | Proportion of women with GDM N (%) |
| --- | --- | --- |
| < 360 nmol/L (deficient) | 7 | 1 (14.3) |
| 360 – 1400 nmol/L (normal) | 498 | 62 (12.5) |
| >1400 <1790 nmol/L (elevated) | 389 | 63 (16.2) |
| ≥1790 nmol/L (excess) | 297 | 55 (18.5) |

GDM prevalence increased progressively across RCF strata in the STOP cohort: from 12.5% among women with normal RCF to 16.2% among women with elevated RCF, and 18.5% among women with excess RCF concentrations (Table 4). Given that one GDM case was observed among only seven women with folate deficiency, the small sample size limits interpretation of this subgroup.

Compared to women whose RCF was within the reference range, those with RCF excess had 48% more GDM cases (p=0.03).

### Placenta but not pituitary secreted hormone concentrations are altered post FA fortification

Numerous hormones regulate maternal insulin resistance and insulin secretion during pregnancy. We investigated whether some of these hormones were altered in women post FA fortification.

Because pregnancy hormone concentrations can fluctuate across gestation (17), we accounted for gestational age at sampling using a linear model with cluster-robust sandwich variance, based on repeated blood sample measures from the same SCOPE participants at 11-13 and 14-

16 weeks’ (N=22). This approach enabled estimation of marginal mean hormone concentrations at 16 weeks’ for both SCOPE and STOP cohorts - a time point that marks the beginning of the exponential rise in hormones and represents the latest sampling time in SCOPE.

Pituitary secreted PRL was not different between SCOPE and STOP. However, compared to SCOPE, placental secreted hPL and GH2 concentrations were 25% and 13% higher in STOP (Table 5).

**Table 5.** Hormone concentrations, presented as Estimated Marginal Means (SEM), at 16 weeks’ gestation for SCOPE and STOP women.

| Estimated Marginal Means (95% CI) | SCOPE | STOP | P |
| --- | --- | --- | --- |
| PRL (ng/ml) | 157.7 (148.7, 167.2) | 145.8 (126.8, 167.65) | 0.3 |
| hPL (µg/ml) | 191.8 (187.2, 196.5) | 239.8 (225.0, 255.5) | <0.0001 |
| GH2 (ng/ml) | 4.7 (4.5, 4.8) | 5.3 (4.9, 5.7) | 0.008 |

SCOPE and STOP sample numbers used for each analysis are shown in Figure 2. Data are adjusted for gestational age at sampling.

Interestingly, hPL as well as PRL concentrations were significantly higher in STOP women with RCF excess compared to those within the normal range - hPL was 12.8% higher (Median [IQ range]: 56.6 [37.2-87.1] vs 63.8 [44.6-97]; p=0.02) and PRL was 24.2% higher (Median [IQ range]: 71.7 [47.4-111.3] vs 88.3 [55.1-133.5]; p=0.005).

### Folate may act via placental hormones to increase risk of GDM

We analysed whether the effect of RCF on GDM involved placental hormones GH2 and hPL. The causal mediation analysis described by Samoilenko *et al.* (2023) (27) was applied. Data were adjusted for maternal age, BMI, ethnicity, SEI, metabolic syndrome and gestational age at sampling. For every 500 nmol/L increase in RCF, GH2 increased and hPL decreased the risk of GDM, which is in line with their established roles in promoting insulin resistance and secretion, respectively. However, given that the two hormones do not act in isolation we implemented the joint mediation analysis described by Vansteelandt *et al.* (2012) (23) (Supplemental Material). There is not enough evidence from this study to show significance of an indirect effect, mediated by GH2 and hPL in combination, on GDM risk (RR [95% CI]: 1.00 [0.99, 1.02]).

## Discussion

Our pregnancy cohort data indicate an alarming tripling in GDM incidence, consistent with the national GDM trajectory (1). Women in both of our pregnancy cohorts were screened for GDM and, importantly, its incidence was calculated using the same WHO diagnostic threshold. It is noteworthy that changes in GDM diagnostic threshold account for an increase in GDM incidence from 4.4 to 5% in SCOPE. This suggests that the change in diagnostic criteria that was implemented in 2015 across Australia is likely to explain some of the increase in national GDM incidence but not the magnitude of the increase nor the steep trajectory. Our study also suggests that known GDM risk factors, such as maternal obesity, age and ethnicity combined cannot completely explain the magnitude of the increase in GDM between the cohorts, as after adjusting data for multiple known confounders, GDM incidence was significantly different between cohorts. Furthermore, other pregnancy complications associated with the same risk factors were not higher in STOP (nor are they rising to the same degree nationally). This suggests that other factors may be contributing to the rising GDM incidence in Australia.

A constellation of known and novel factors has likely contributed to increased national GDM incidence. Based on the data presented herein and supported by evidence from clinical (3–6) and animal studies (7–10), we propose that elevated maternal folate status is a contributing factor to the rising incidence of GDM in Australia. A combination of FA food fortification, both mandatory and voluntary, increased dose of supplementation and continued supplementation beyond the recommended first trimester, have resulted in maternal folate excess, levels that far exceed the established clinical reference range. Previous studies have reported a U-shaped relationship between the dose and duration of FA supplementation and GDM risk (4) (28), and from that perspective excess maternal folate may biologically mimic folate deficiency, as previously reported for other biological measures. Although numbers in the deficient RCF category were low in our cohort, our data also suggest a potential U-shaped association between maternal RCF levels and GDM incidence. Given that detrimental effects of maternal folate deficiency (characterised using similar reference range) on pregnancy health are well documented (20) (29), the lack of evidence for the safety of excess maternal folate is concerning, particularly as folate excess is now far more common than folate deficiency. A recent NIH workshop highlighted the need for comprehensive research to bridge the knowledge gaps in understanding the metabolic and clinical effects of excess folates/FA (30). Addressing this need, we show that increased RCF levels in women post FA fortification significantly increased GDM risk and that women with RCF excess had the highest proportion of GDM. These findings support previous reports of an association between increasing RCF during pregnancy and increased GDM risk (31) (32).

A likely explanation for the observed association between RCF and GDM risk, but the absence of a similar association with serum folate, is the use of non-fasting blood samples in this study. Serum folate reflects recent folate/FA intake and is subject to short-term fluctuations influenced by meal timing and supplement use (33). In contrast, RCF provides a more stable measure of long-term folate status (26), making it a more reliable marker when assessing associations between non-fasting blood samples with pregnancy outcomes such as GDM.

Despite convincing evidence from observational and animal studies for the role of FA in GDM aetiology, to date the mechanisms remain unknown. Our study shows that women with folate excess had higher hPL and PRL (both primarily promote insulin secretion, although hPL has also been reported to promote insulin resistance) compared to women whose folate was within the normal range. Whilst it is possible that higher hPL and PRL levels in women with folate excess may indicate increased glucose tolerance, based on previous work they are more likely to reflect reduced glucose tolerance (34). Whatever the biological relevance of these hormones may be as it relates to hyperglycaemia, the fact that these hormones are altered with maternal folate excess warrants further investigation.

Importantly, our study shows that pregnant women post FA fortification had increased placenta-secreted (hPL and GH2) but not maternal-secreted (PRL) hormones in early gestation. Due to the established roles of these hormones in regulation of maternal glucose homeostasis this finding supports our hypothesis that the placenta may be involved. Whether the observed indirect effect of maternal folate on GDM via individual placental hormones has biological relevance remains to be determined. The observation that increased maternal folate acts via GH2 and hPL to increase and reduce GDM risk, respectively, coheres with what we know about the roles of these hormones in promoting insulin resistance and secretion, respectively (17) (18) (35). Lack of evidence for the indirect effect of maternal folate on GDM via placental hormones using *combined* hormone mediation analysis could be due to a combination of factors: 1) insufficient samples from this cross-sectional study; 2) the effects of these two specific hormones cancel each other systemically; 3) additional hormones that we are yet to analyse are also involved. The role of the placenta in mediating the effects of FA on GDM needs to be explored for all placental peptides (including leptin, oestradiol, progesterone etc) known to regulate maternal glucose homeostasis. Other than our recently published study which showed alterations in placental hormones in women with uncomplicated pregnancies following FA fortification (13), we are unaware of other studies that have investigated the relationship between FA and placental hormones in the context of insulin resistance, hyperglycaemia and/or GDM. Further studies are warranted to determine why some women may be more sensitive than others to the effects of FA in the context of GDM aetiology.

In conclusion, our study confirms a significant increase in GDM incidence in women post FA fortification and we show that increased maternal folate status in early gestation is a novel independent risk factor for GDM that has likely contributed to the national GDM rise. Further investigation is warranted to establish the role of FA in the regulation of placental endocrine function to promote insulin resistance. Urgent assessment of the safety of maternal folate excess in pregnancy is required. Evaluations of efficacy and safety of the current real world FA supplementation practice during pregnancy in the era of FA food fortification are warranted. Improved guidelines on FA supplementation during pregnancy would protect the foetus in early gestation against NTDs but also protect the mother and foetus from adverse effects of hyperglycaemia. Given the duration and extent of FA supplementation in pregnancy is highly modifiable, understanding potential harms of excess FA intake is of major public health importance.

## Data Availability

Data that support the findings of this study are openly available in Flinders University Repository of Open Access Data Sets (ROADS) at http://doi.org/10.25451/flinders.21721397, reference number 10.25451/flinders.21721397. All protocols and models related to this project are available upon reasonable written request to the corresponding authors.

## Acknowledgements

The authors would like to extend their gratitude to all the women who participated in SCOPE and STOP. We also thank the research staff involved in recruitment and collection of biological samples from the participants (SCOPE: Denise Healy, Karen Rivers, Jessica Phillips; STOP: Julia Dalton, Samantha Paul, Petra Verburg, Jessica Phillips, Suzette Coat, Caitlin McCullough).

## Funding and Assistance

This study was made possible due to funding from the National Health and Medical Research Council Investigator (GNT1174971; Awarded to CTR), National Health and Medical Research Council Project (GNT1161079; Awarded to CTR, GAD and SL) and Flinders Foundation Health Seed (Awarded to TJK, CTR, SL and AA) grants. CTR was supported by the Matthew Flinders Fellowship.

## Conflicts of Interest

The authors declare no conflict of interest.

## Data Sharing Statement

Data that support the findings of this study are openly available in Flinders University Repository of Open Access Data Sets (ROADS) at http://doi.org/10.25451/flinders.21721397 reference number 10.25451/flinders.21721397. All protocols and models related to this project are available upon reasonable written request to the corresponding author.

## Author Contributions and Guarantor Statement

CTR and GAD conceptualized and established the SCOPE (Adelaide) and STOP cohorts. GAD was involved in patient recruitment in both SCOPE and STOP studies. DM was involved in biological sample handling and biobanking for both cohorts. TJK, GAD and CTR conceptualized the study reported. TJK, DM, ALA and JW conducted the laboratory experiments. TJK, SL, MM and MDS analysed the data. TJK wrote the manuscript with input from other authors. All authors approved the final version of the manuscript. T.J-K. and S.L. are the guarantors of this work and, as such, had full access to all the data in the study and take responsibility for the integrity and the accuracy of the data analysis.

## Notes

### Competing Interest Statement

The authors have declared no competing interest.

### Author Declarations

Central Northern Adelaide Health Service Human Research Ethics Committee (REC1712/5/2008) and the Womens and Childrens Health Network Human Research Ethics Committee (HREC/14/WCHN/90) gave ethical approval for this work. SCOPE and STOP were registered with the Australian New Zealand Clinical Trial Registry (ACTRN12607000551493 and ACTRN12614000985684, respectively).

