## Supplemental Data for "Folate Overload and the Placental Hormone Axis: A Hidden Risk for Gestational Diabetes Mellitus"

**Supplementary Figure 1. IPTW analysis assumptions**

The direct acyclic graph (DAG) for the individual participant data meta-analysis is as follows:

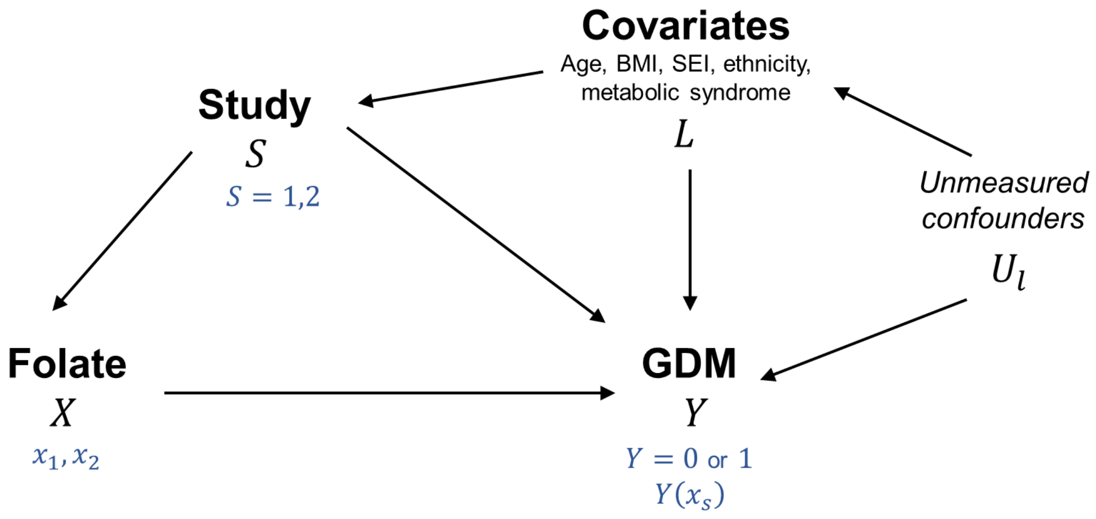


The following assumptions are made:

1) Consistency:

   The counterfactual outcome ($Y(x_s)$) agrees with the observed outcome $Y$ for all individuals in study ($S$) exposed to folate ($X$). It is assumed that folate ($X$) is the only data generating mechanism for GDM ($Y$) and that the counterfactual outcome can be predicted from the observed $X$ and $Y$.

2) Positivity: $f(X = x|L) > 0$ and $0 < P(S_i = j|L_i) > 1$

   There is balanced confounding between folate exposure status, such that any individual with characteristics ($L$) has a positive probability of being included (i.e. eligible to participate) in the other study, and positive probability of exposure to folate.

3) Ignorable treatment assignment: $Y(x_s) \perp X|L$

   Within each study, folate exposure ($X$) status is independent of the counterfactual outcome ($Y(x_s)$), conditioning on covariates ($L$). This assumes that individuals within a study with the same characteristics ($L$) would have the same outcome risk if given the same folate exposure.

4) Ignorable study assignment: $Y(x_s) \perp S|L$

   Study indicator ($S$) is independent of the counterfactual outcome ($Y(x_s)$), conditioning on covariates ($L$). This assumes that individuals in different studies with the same characteristics ($L$) would have the same outcome risk if given the same folate exposure.

**Supplementary Figure 2. Direct acyclic graph (DAG) for mediation analysis**

Direct effect of maternal red cell folate on GDM is shown with black arrow. Blue arrows indicate the joint indirect effect of folate on GDM through hPL and GH2 (Folate → hPL→ GDM; folate → GH2 → GDM; folate → hPL → GH2 → GDM).

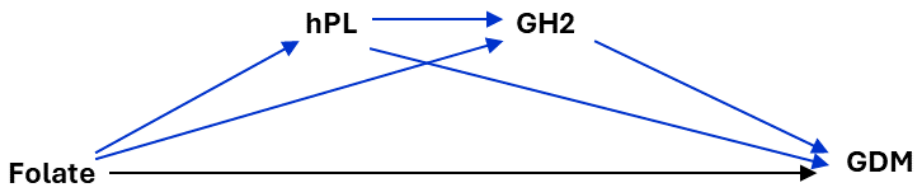
